# Imaging and Motor Asymmetry in Parkinson’s Disease associated with GBA and LRRK2 mutations: Data from the PPMI study

**DOI:** 10.64898/2026.11.17.26368329

**Authors:** Konstantinos Patsantzopoulos, Nikolaos Papagiannakis, Christos Koros, Leonidas Stefanis, The Parkinson’s Progression Markers Initiative

**Affiliations:** Department of Neurology, Eginition Hospital, Medical School, National and Kapodistrian University of Athens, Athens, Greece

**Author notes:** Corresponding author, 1st Department of Neurology, Eginition Hospital, Medical School, National and Kapodistrian, University of Athens, Athens, Greece, 72-74 Vas. Sofias Ave, Athens 11528, Greece. The author list for the Parkinson’s Progression Markers Initiative can be found in the appendix.

## Abstract

**Background:** According to the α-Synuclein Origin site and Connectome (SOC) model, idiopathic Parkinson’s Disease (iPD) is classified into two distinct subtypes: symmetric body-first, with diffuse spread and early non-motor clinical manifestations, and asymmetric brain-first, with initially circumscribed motor manifestations. Evidence regarding the application of this model to genetic PD remains limited.

**Objective:** To investigate imaging and motor asymmetry in genetic vs iPD and idiopathic REM Sleep Behavior Disorder (iRBD), addressing thus indirectly the SOC model in genetic PD.

**Methods:** We analyzed demographic, imaging and clinical data from 85 GBA PD, 166 LRRK2 PD, and 912 iPD patients, as well as 30 iRBD participants with abnormal DATSCAN from the PPMI database. Imaging asymmetry was assessed both by Asymmetry Index and absolute Asymmetry difference. Motor asymmetry was calculated as the absolute difference between the MDS-UPDRS III scores of the right and left sides.

**Results:** GBA- and LRRK2-PD patients exhibit more symmetric putaminal radioligand uptake than iPD patients, but greater imaging asymmetry than iRBD subjects in the caudate. iRBD is more symmetric than iPD in both caudate and putamen. Consistent with the imaging data, GBA- and LRRK2-PD exhibit greater clinical motor symmetry than iPD.

**Conclusion:** GBA- and LRRK2-PD are more symmetric than iPD on imaging and clinical grounds. GBA-PD appears to be a predominantly symmetric diffuse synucleinopathy, as predicted by the SOC model. The SOC model may not be applicable to LRRK2-PD, with its predominantly circumscribed motor phenotype, possibly because in this case α-Synuclein pathology does not determine lateralization of motor manifestations.

## Introduction

Parkinson’s disease (PD) is a heterogeneous disorder characterized by a variable spectrum of motor and non-motor symptoms, differing in both severity and progression rate among individuals. The diagnosis of PD is mainly clinical and relies on the presence of core motor symptoms, including bradykinesia, rigidity, and/or resting tremor, typically affecting one side of the body at the beginning of the disease. DATSCAN imaging, a dopamine transporter (DAT) single-photon emission computed tomography (SPECT) technique, is used as supportive evidence for PD diagnosis and as a research tool. The radioligand uptake in DATSCAN is generally reduced in the striatum contralateral to the side of the body displaying more severe motor symptoms, reflecting the underlying asymmetric neurodegeneration.

While unilateral motor symptom onset is recognized as a supportive diagnostic criterion for PD, it is notable that approximately 30-40% of patients exhibit relatively symmetric motor symptoms at the time of diagnosis [1-6]. The concept of imaging and motor asymmetry in PD has been studied over the years, yielding diverse perspectives on its underlying pathophysiological mechanisms and prognostic implications. Age at disease onset appears to be significantly correlated with both imaging and motor asymmetry. According to the age at which the initial symptoms manifest, PD can be categorized into early-onset (EOPD) and mid or late-onset PD (M-LOPD), with most studies adopting 50 years as the threshold distinguishing the two subtypes. We have shown that EOPD is associated with greater striatal asymmetry on DATSCAN imaging, a difference that tends to persist over time, particularly within the caudate nucleus [7]. These findings are further supported by evidence showing that a more symmetric presentation of motor symptoms is observed at a higher frequency among individuals with M-LOPD [8, 9]. In addition to classifying PD by age at disease onset, multiple efforts have been undertaken to delineate distinct subtypes within the PD population, aiming to provide a clearer understanding of the disorder’s heterogeneous nature. Notably, PD can be categorized into tremor-dominant (tPD) and akinetic-rigid (arPD) subtypes based on the predominant motor symptoms at disease onset. Although imaging asymmetry using DATSCAN has been examined between these subtypes, no statistically significant differences have been identified [10]. To further specify clinical phenotypes within the PD population, cluster analysis has been employed to classify patients into distinct subgroups characterized by their symptom profiles and progression patterns. In one such study, three disease clusters were identified: a motor/slow progression cluster, an intermediate cluster, and a diffuse/malignant disease cluster. Patients within the diffuse/malignant group generally exhibited severe motor and non-motor symptoms and showed lower asymmetry scores, with fewer patients experiencing unilateral motor symptom onset [11].

Recently, the α-Synuclein Origin site and Connectome Model (SOC model) has been proposed to provide a biologically grounded explanation for the observed heterogeneity in motor and imaging asymmetry [12]. This model distinguishes Parkinson’s disease into two subtypes based on where α-synuclein pathology begins and how it spreads: the body-first subtype, originating in the enteric nervous system and advancing to the brain via the vagus nerves, leading to a more symmetric involvement of the substantia nigra and the brain-first subtype, beginning in the central nervous system and spreading asymmetrically. Patients presenting with REM sleep behavior disorder (RBD) preceding the onset of motor symptoms are classified as body-first, whereas individuals who either do not exhibit RBD or in whom RBD emerges after motor symptom onset are categorized as brain-first. Multiple investigations have explored the association between RBD status and DATSCAN asymmetry, with several reporting that the premotor occurrence of RBD correlates with a more symmetric reduction in radioligand uptake on DATSCAN imaging [13, 14].

The SOC model mainly applies to idiopathic PD, as evidence for its relevance to genetic forms remains limited. Familial PD constitutes approximately 5-10% of all PD cases, with over 20 distinct PARK gene loci implicated in its Mendelian inheritance [15,16]. Pathogenic variants in the leucine-rich repeat kinase 2 (*LRRK2*) gene and in the glucosylceramidase beta (*GBA1*) gene are the two most common genetic risk factors for PD, accounting for up to 10% of cases worldwide. A limited number of studies have addressed the issue of motor and imaging asymmetry in these genetic forms of PD. GBA-PD has been associated with a higher frequency of symmetric onset of motor symptoms in comparison to idiopathic PD [17,18], even though DATSCAN imaging has shown no significant differences in the asymmetry of radiotracer uptake between these two groups [19,20]; likewise, no differences in DATSCAN asymmetry were reported in LRRK2-PD compared to iPD [20, 21].

In an effort to disentangle the issue, we sought to investigate rigorously the issue of asymmetry in genetic vs iPD both clinically, based on motor asymmetry on MDS-UPDRS III examination, and on imaging, by evaluating Striatal Binding Ratios (SBRs) on DATSCAN, using different methods to assess asymmetry, in the large Parkinson’s Precision Medicine Initiative (PPMI) cohort. We included additionally from the same cohort iRBD subjects with abnormal DATSCANs, by definition evolving into “body-first PD”, to also examine potential differences on DATSCAN SBR asymmetry between this group and genetic PD, something that has not been previously performed. In this way, using asymmetry as a proxy, we hoped to indirectly address the issue of the SOC model in the context of genetic PD.

## Methods

### Data Acquisition and Sharing

Data used in the preparation of this article were obtained on July 14, 2025, from the PPMI database (https://www.ppmi-info.org/access-data-specimens/download-data; RRID:SCR_006431). Data considering prodromal patients were part of tier 3 data availability. For up-to-date information on the study and data availability, visit http://www.ppmi-info.org. Besides DATSCAN measurements, basic demographic characteristics (age at visit, age at onset, and disease duration), and basic clinical scores (Unified Parkinson’s Disease Rating Scale [UPDRS] Part III) were also collected.

### Participants

We have included all participants participating in the Parkinson’s Disease PPMI cohort harboring mutations in *GBA1* and *LRRK2* genes, as well as participants without any known PD-associated pathogenic variants that were not excluded or screen failed. In addition, participants in the prodromal cohort with RBD were included if their DATSCAN was visually interpreted as indicative of neurodegeneration. Only data from the screening visit, for DATSCAN and basic demographic characteristics, and baseline visit, for clinical data, were utilized.

### Asymmetry Measures

An asymmetry index for DaTSCAN was computed from semiquantitative striatal binding ratios (SBRs) in homologous regions of interest (ROIs) in each hemisphere. For each subject, left and right SBRs were derived for the caudate and putamen using standardized ROIs normalized to an occipital reference region, and an asymmetry index (AI) was calculated as:

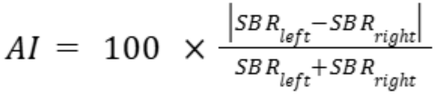

The asymmetry difference was calculated by using the absolute value of the SBR difference between the left and right side.

Both Asymmetry Index and asymmetry difference were computed separately for caudate and putamen.

The motor asymmetry was quantified as the absolute difference between the sum of right- and left-sided scores for UPDRS Part III (Motor Examination) items scored separately for each side. Specifically, the sum of items 3.3b, 3.3d, 3.4-3.8a, 3.17c and 3.15-3.17a was subtracted from the sum of items 3.3c, 3.3e, 3.4-3.8b, 3.17b and 3.15-3.17b and then the absolute value of the result was calculated. Items assessing axonal signs were not evaluated.

### Statistical Analysis

Statistical analysis was performed using R version 4.5. All data were tested for normality using the Shapiro–Wilk test and were determined to be non-normally distributed. The Kruskal–Wallis test was applied, followed by Dunn’s post hoc test for pairwise comparisons, to evaluate differences across the various groups.

Data is presented as median (interquartile range) in the tables and text. Motor asymmetry was calculated as the absolute difference between the MDS-UPDRS part III items corresponding to the left and right sides, excluding items related to axonal features.

An ANCOVA model incorporating time from diagnosis was employed to adjust p-values for disease progression. All presented p-values comparing disease groups are duration-adjusted.

The statistical significance limit was set at p = 0.05.

## Results

### Demographic and clinical characteristics

Demographic and clinical characteristics are shown in Table 1. We analysed 85 GBA-PD, 166 LRRK2-PD, 912 iPD patients and 30 iRBD participants with abnormal DATSCAN. The iRBD cohort had a higher mean age at assessment than the PD groups. Patients with GBA- and LRRK2-associated PD had longer disease duration since diagnosis than iPD, and GBA-PD patients had the highest MDS-UPDRS III scores, followed by iPD and then LRRK2-PD. Of the GBA-PD participants 75 (83%) were on dopaminergic medication, 141 (84%) of the LRRK2-PD and 846 (92%) of the iPD.

**Table 1.** Demographic and clinical characteristics of patient groups. p values are adjusted for disease duration where applicable. Data is presented as median (interquartile range)

|  | GBA-PD | LRRK2-PD | iPD | RBD | p-value |
|---|---|---|---|---|---|
| Sample size (N) | 85 | 166 | 912 | 30 |  |
| Sex (F/M) | 41/44 | 83/83 | 322/590 | 6/24 | <0.001 |
| Age at visit (years) | 64.1 (57.8 – 71.3) | 65.7 (57.6 – 71.4) | 63.8 (56.7 – 69.7) | 70.05 (66.73 – 74.3) | <0.001 |
| Disease duration since diagnosis (months) | 26 (12 – 50) | 26 (12 – 52) | 4 (2 – 9) |  | <0.001 |
| UPDRS-III | 24 (15 – 31) | 17 (13 – 26) | 19 (14 – 25) |  | 0.012 |

### DATSCAN Asymmetry Asymmetry Index

The asymmetry index (AI) data are presented in Table 2, and Figure 1. **Caudate AI** was 8.56 in iPD, 8.91 in GBA-PD, 7.8 in LRRK2-PD and 4.69 in iRBD. There were no significant differences between GBA-PD, LRRK2-PD and iPD. In contrast, iPD, GBA-PD and LRRK2-PD all had higher caudate AI than iRBD (p<0.001, p=0.001 and p=0.007 respectively). **Putaminal AI** was 7.18 in iPD, 5.22 in GBA-PD, 5.56 in LRRK2-PD and 4.86 in iRBD. (figure 1). iPD had higher putamen AI than GBA-PD and iRBD (p=0.007 for both), as well as LRRK2-PD (p=0.025). There was no difference in putaminal AI between GBA-PD, LRRK2-PD and RBD.

**Table 2.**
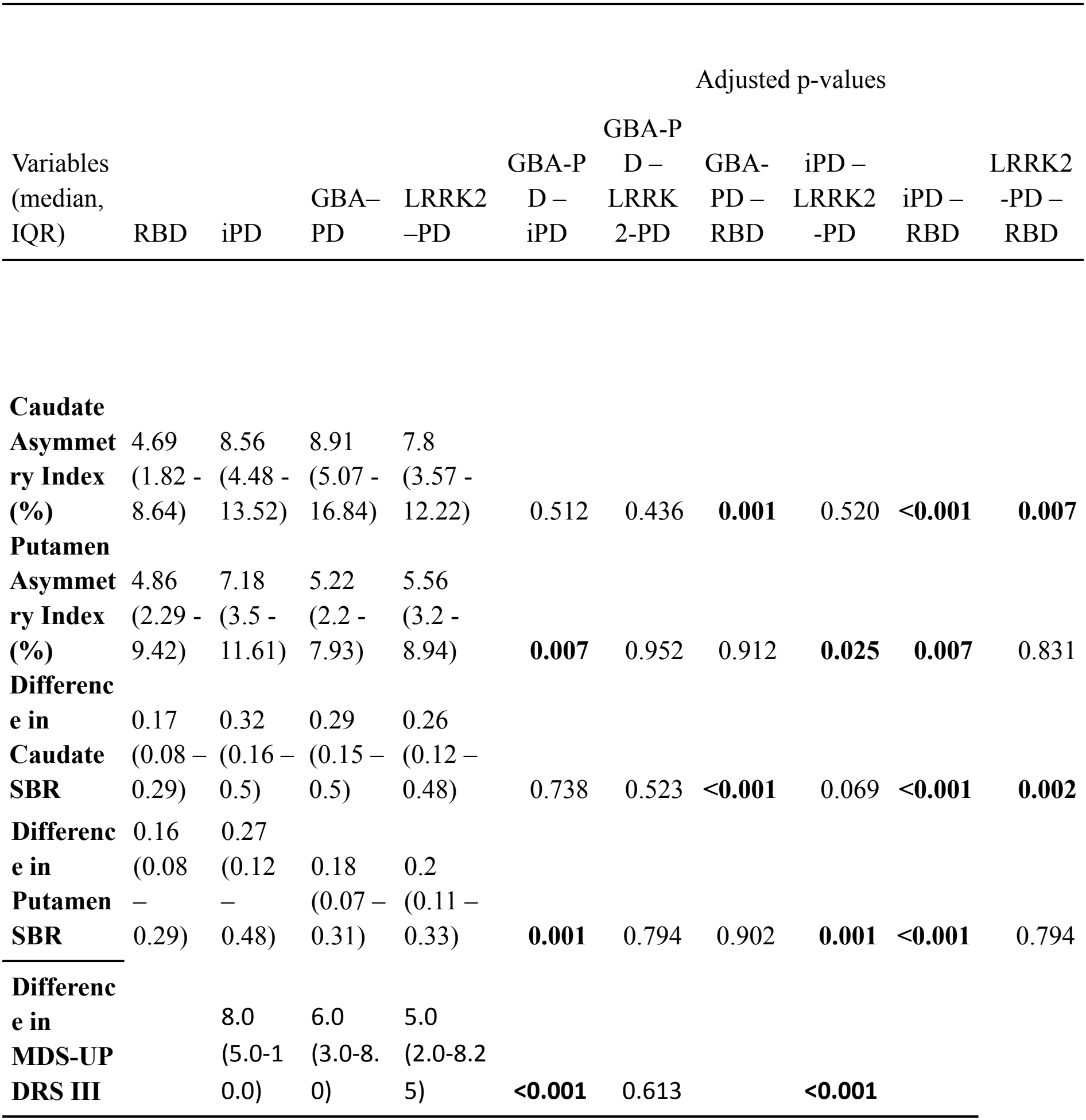
Summary of asymmetry measurements in the four patient groups (supplementary material). p values are adjusted for disease duration where applicable.Data is presented as median (interquartile range).

|  |  |  |  |  | **Adjusted p-values** |  |  |  |  |  |
|---|---|---|---|---|---|---|---|---|---|---|
| Variables (median, IQR) | RBD | iPD | GBA–PD | LRRK2–PD | GBA-PD – iPD | GBA-PD – LRRK2-PD | GBA-PD – RBD | iPD – LRRK2-PD | iPD – RBD | LRRK2-PD – RBD |
| **Caudate Asymmetry Index (%)** | 4.69 (1.82 - 8.64) | 8.56 (4.48 - 13.52) | 8.91 (5.07 - 16.84) | 7.8 (3.57 - 12.22) | 0.512 | 0.436 | **0.001** | 0.520 | **<0.001** | **0.007** |
| **Putamen Asymmetry Index (%)** | 4.86 (2.29 - 9.42) | 7.18 (3.5 - 11.61) | 5.22 (2.2 - 7.93) | 5.56 (3.2 - 8.94) | **0.007** | 0.952 | 0.912 | **0.025** | **0.007** | 0.831 |
| **Difference in Caudate SBR** | 0.17 (0.08 – 0.29) | 0.32 (0.16 – 0.5) | 0.29 (0.15 – 0.5) | 0.26 (0.12 – 0.48) | 0.738 | 0.523 | **<0.001** | 0.069 | **<0.001** | **0.002** |
| **Difference in Putamen SBR** | 0.16 (0.08 – 0.29) | 0.27 (0.12 – 0.48) | 0.18 (0.07 – 0.31) | 0.2 (0.11 – 0.33) | **0.001** | 0.794 | 0.902 | **0.001** | **<0.001** | 0.794 |
| **Difference in MDS-UPDRS III** |  | 8.0 (5.0-10.0) | 6.0 (3.0-8.0) | 5.0 (2.0-8.25) | **<0.001** | 0.613 |  | **<0.001** |  |  |

**Figure 1.**
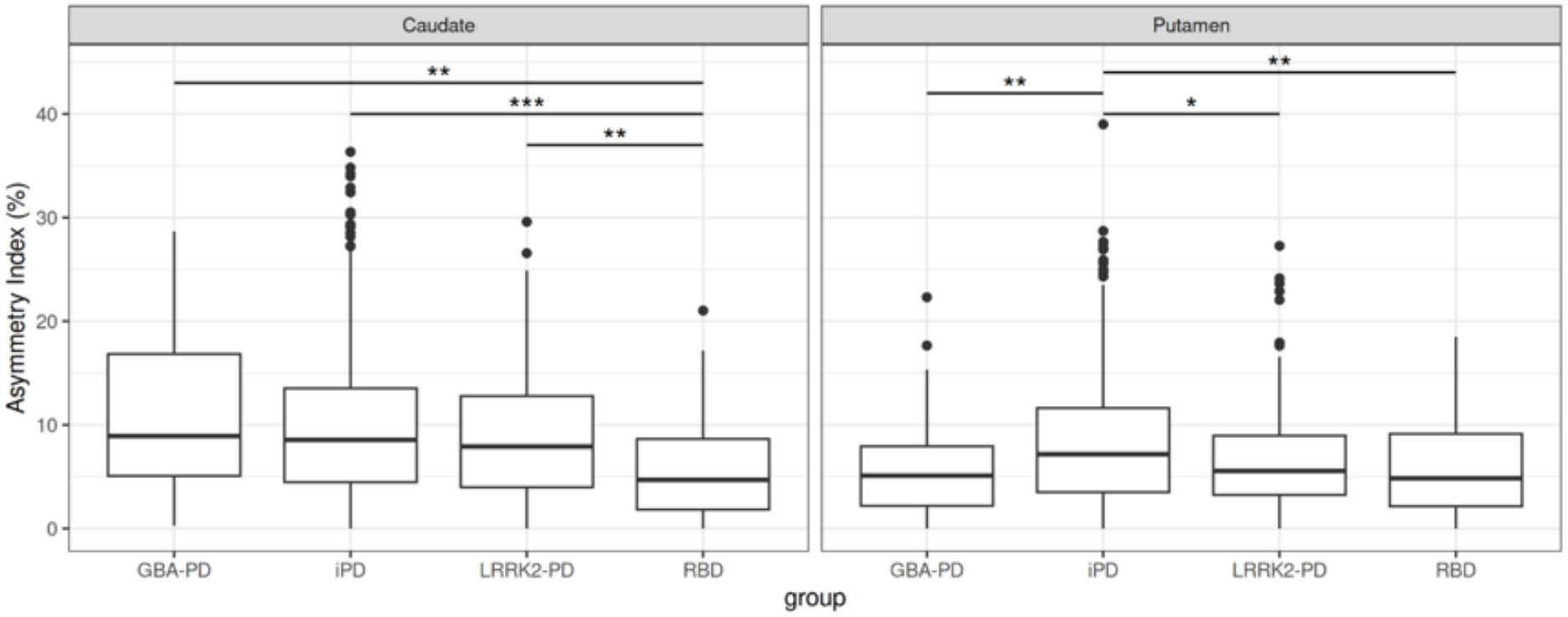
Asymmetry Index. Caudate nucleus and putaminal DAT Asymmetry Indices plotted per group (The central line represents the **median**, and the box spans the 25th–75th percentiles (interquartile range), containing the middle 50% of the data). **p values are adjusted for disease duration where applicable**.

### SBR difference

Using the absolute right–left SBR difference, **caudate asymmetry** was 0.32 in iPD, 0.29 in GBA-PD, 0.26 in LRRK2-PD and 0.17 in iRBD (Table 2). GBA-PD and LRRK2-PD did not differ from iPD (p=0.738 and p=0.069), but they had greater caudate asymmetry than iRBD (p<0.001 and p=0.002), whereas iPD had markedly greater caudate asymmetry than iRBD (p<0.001) (figure 2). **Putamen asymmetry** was 0.27 in iPD, 0.18 in GBA-PD, 0.20 in LRRK2-PD and 0.16 in iRBD (Table 2). iPD showed greater putamen asymmetry than GBA-PD, LRRK2-PD and iRBD (p=0.001 for all). There was no difference in putamen asymmetry between GBA-PD, LRRK2-PD and iRBD (figure 2).

**Figure 2.**
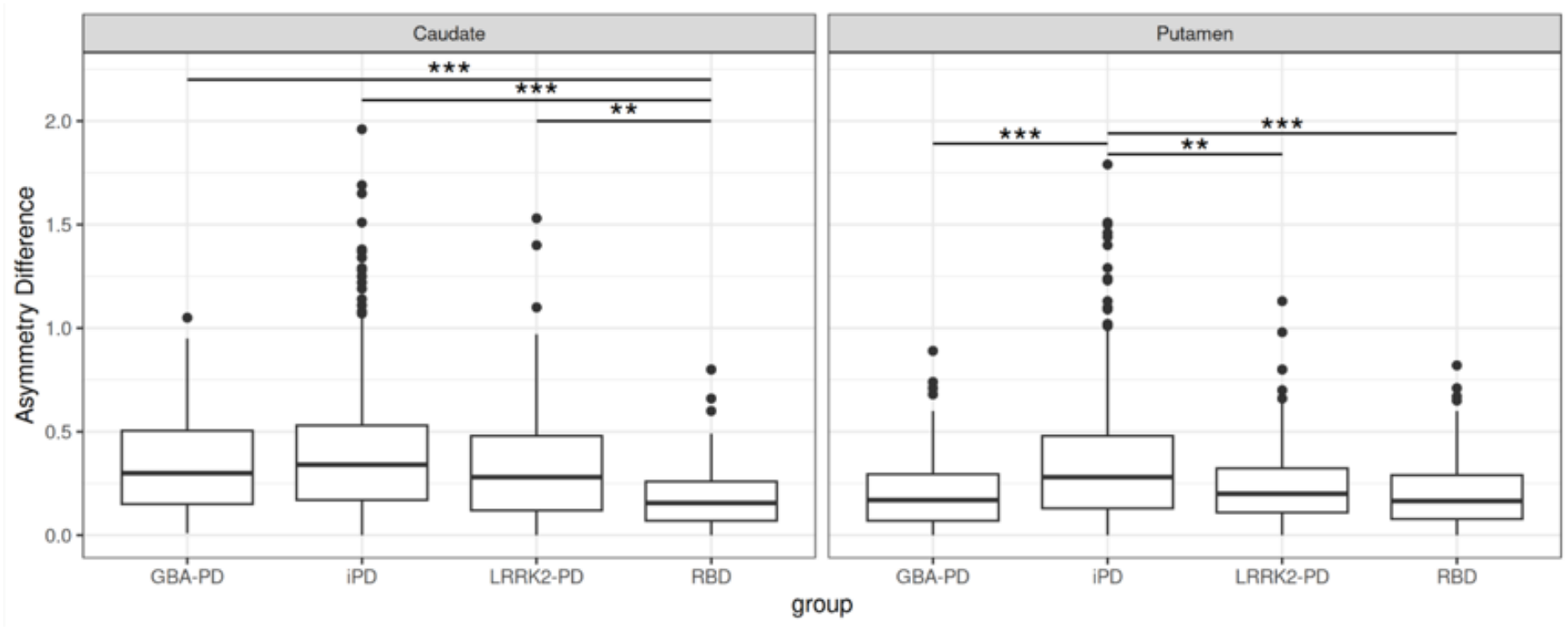
Asymmetry Difference. Absolute difference between right and left caudate nucleus and putamen SBR plotted per group (The central line represents the **median**, and the box spans the 25th–75th percentiles (interquartile range), containing the middle 50% of the data). **p values are adjusted for disease duration where applicable**.

### Motor Asymmetry

Motor asymmetry, defined as the absolute right–left MDS-UPDRS III difference, was lower in GBA-PD and LRRK2-PD than in iPD (Figure 3). Median values were 8.0 for iPD, 6.0 for GBA-PD (p<0.001 vs iPD) and 5.0 for LRRK2-PD (p<0.001 vs iPD). GBA-PD and LRRK2-PD did not differ between them.

**Figure 3.**
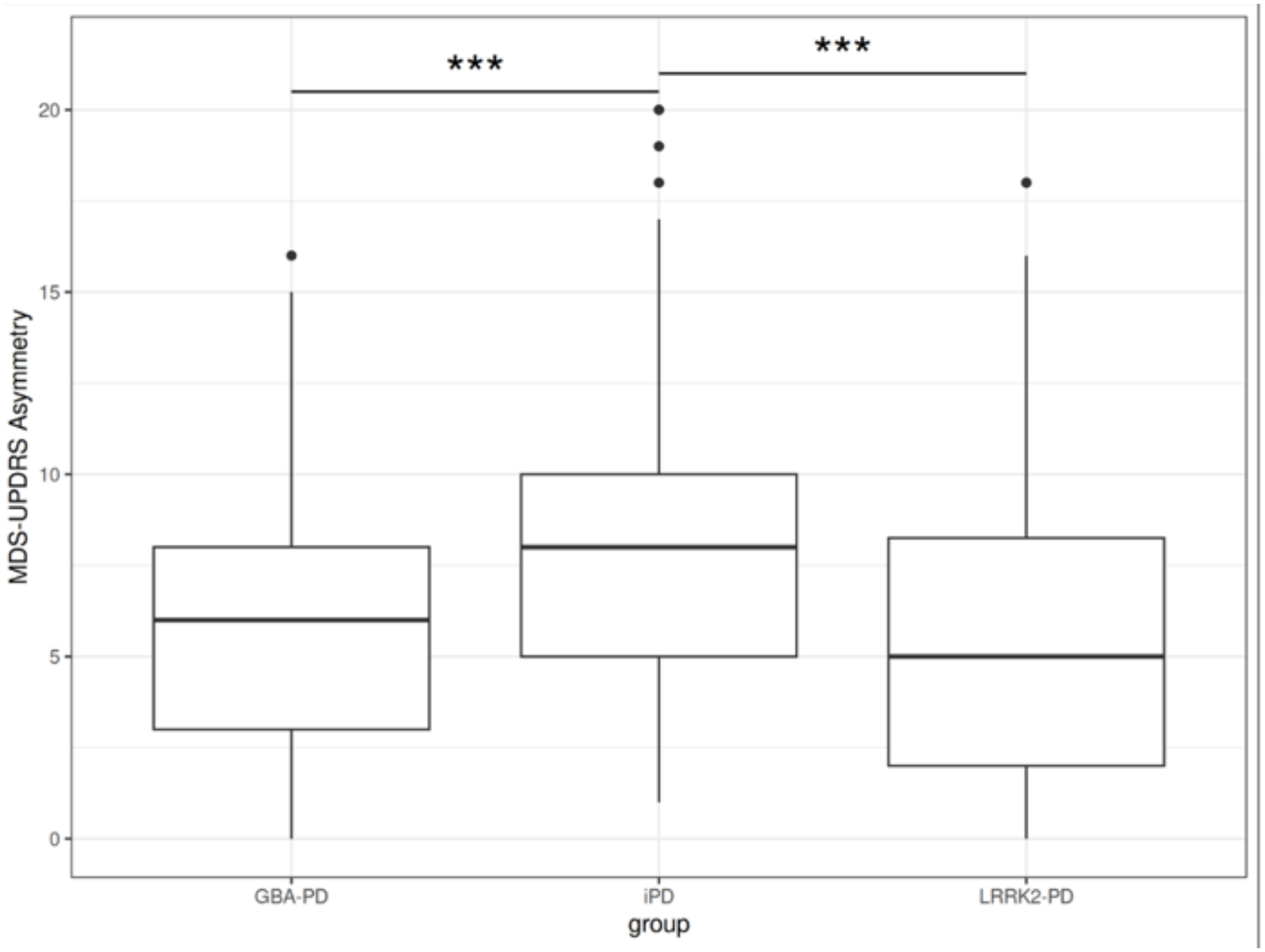
Motor Asymmetry. Absolute difference between right and left scores of the UPDRS-III scale in idiopathic Parkinson’s disease (iPD), GBA- and LRRK2-associated PD patients (The central line represents the **median**, and the box spans the 25th–75th percentiles (interquartile range), containing the middle 50% of the data). **p values are adjusted for disease duration where applicable.**

## Discussion

Employing diverse metrics to assess nigrostriatal dysfunction asymmetry, we studied a total of 85 GBA-PD, 166 LRRK2-PD, 912 iPD patients and 30 iRBD subjects with abnormal DATSCAN from the PPMI cohort. We demonstrated that GBA and LRRK2-PD patients exhibited greater imaging asymmetry than iRBD subjects in the caudate nucleus, while displaying lower putaminal asymmetry than iPD by the asymmetry-index and absolute-difference measures. Motor asymmetry metrics aligned with putaminal imaging findings. Related to our results, McNeill et al. reported that GBA- and LRRK2-PD are linked to relatively asymmetric striatal dopaminergic loss, but they compared them with healthy controls, not iPD or iRBD patients[22].

The SOC model proposes that phenotypic heterogeneity in PD is determined by the site of initial α-synuclein aggregation and its subsequent propagation along anatomically defined neural networks [12]. According to this framework, PD can be divided into a body-first subtype, where pathology starts in the enteric or autonomic nervous system and spreads symmetrically to the brain, leading to symmetric substantia nigra involvement and characterized by prominent non-motor symptoms, including early prodromal; and a brain-first subtype, where pathology originates in the CNS (e.g., amygdala or olfactory bulb) and spreads predominantly ipsilaterally, resulting in greater motor asymmetry without prominent early non-motor symptoms. In the body-first subtype, caudo-rostral spread involves pontine nuclei implicated in REM sleep behavior disorder (RBD), such that RBD typically precedes motor onset, whereas in the brain-first subtype RBD is absent or occurs after motor symptom onset. This is supported by Knudsen et al., who showed more symmetric striatal degeneration in de novo PD with premotor RBD versus without. [13] However, Lovdal et al. found no DATSCAN or motor asymmetry differences between PD-RBD+ and PD-RBD-, thereby challenging the direct clinicopathological correlation proposed by the SOC framework [23]; it has to be noted however that this study did not distinguish these categories at disease onset.

Based on the SOC model, iPD patients represent a mixed group of body-first and brain-first PD, whereas iRBD subjects comprise a pure group of body-first PD. This would suggest that iPD would be more asymmetric than iRBD, a finding present in earlier studies [13], which was confirmed here. GBA-PD and LRRK2-PD stand in the middle of this continuum, embodying a distinct subgroup that exhibits greater DATSCAN asymmetry than the pure body-first subgroup of iRBD and a more symmetric radioligand uptake compared to the mixed iPD group. Relative putaminal symmetry in these genetic forms of PD compared to iPD is compatible with our findings regarding the clinical index of asymmetry. To our knowledge, this is the first study that has combined assessment of DAT asymmetry with that of clinical motor asymmetry. It is especially noteworthy that these separate assessments converge on the same conclusion, that these forms of genetic PD are more symmetric than iPD.

Based on the phenotype of GBA-PD, which is more severe compared to iPD and includes more prominent non-motor symptoms such as dementia and RBD [24, 25], the initial hypothesis was that it would be very similar, in terms of asymmetry indices, to the body-first iRBD; this was partially borne out, as putaminal asymmetry was less pronounced compared to the presumed mixed group of iPD, and was not different from iRBD; however caudate asymmetry in GBA-PD was more pronounced than the body-first iRBD and similar to iPD. On the other hand, LRRK2-PD tends to be more motor-centered, with less non-motor symptoms compared to iPD [26], and this led to the initial prediction that it would be even more asymmetric than the mixed group of iPD; however this was clearly not the case, in fact the opposite occurred. These results may indicate that, in the LRRK2-PD group at least, alpha-synuclein spread may not be a major determinant of pathogenicity and lateralization of motor manifestations.

In contrast to our work, Jeong et al., using a smaller PPMI cohort, did not demonstrate significant differences in asymmetry indices between iPD, GBA-PD and LRRK2-PD. [20] This discrepancy may be explained by both differences in sample size and methodological variations between the two studies. The smaller sample size in the study by Jeong et al. may have limited the statistical power to detect significant between-group differences. Furthermore, methodological differences, particularly the time point of the DATSCAN images utilized for the iPD cohort, may have contributed to the divergent findings. Specifically, Jeong et al. based their analysis on the two-year follow-up data of the iPD cohort and utilized DATSCAN images acquired at the two-year follow-up visit, whereas we analysed DATSCAN images obtained at the screening visit. Several studies have demonstrated that, as PD progresses, neurodegeneration increasingly involves both striata, leading to greater symmetry over time [27-29]. Consequently, potential group specific differences in asymmetry may have been obscured at the later stage of disease progression assessed by Jeong et al. Additional studies involving small cohorts of genetically defined PD patients have similarly failed to demonstrate significant differences in DATSCAN asymmetry [19, 21]. We should note that our work here reports the largest sample size used to assess this issue. Furthermore, different types of gene mutations, ranging from mild to severe in their pathogenic effects, may influence both the pattern of disease manifestations and the rate of neurodegeneration, thereby affecting the symmetry of radioligand uptake.

This study has several strengths. A major one is that all demographic, imaging, and clinical data were obtained from the PPMI study, an international, multicenter, observational cohort study that longitudinally collects and regularly updates biological and clinical data. The use of a single, well-characterized data source enhances data harmonization, facilitates integrated analyses, and strengthens the overall validity and credibility of the findings. Additionally, the large sample size of GBA-PD and LRRK2-PD cohorts, combined with the simultaneous evaluation of both imaging and clinical measures of asymmetry, represents a key novel aspect of this study. Importantly, adjusting p-values for disease duration mitigates potential bias arising from the tendency of PD to become more symmetric as the disease progresses.

Despite these strengths, several limitations should be acknowledged. A major limitation in the study of PD asymmetry is the lack of standardized metrics for quantifying imaging and motor asymmetry. Different research groups employ heterogeneous approaches, which hampers direct comparison across studies and limits the generalizability of the conclusions. Moreover, the DATSCAN asymmetry index (AI) represents a nonlinear, relative measure that exhibits exponential growth with lower SBR values; therefore, absolute SBR differences may constitute a more appropriate metric for the assessment of asymmetry. It is important that in the present study both measures of DAT asymmetry, assessed independently, gave similar results. In addition, each genetic PD subgroup comprises a heterogeneous spectrum of gene mutations, which may differentially influence phenotypic expression. The relatively small sample size of individuals with iRBD and abnormal DATSCAN findings reduces statistical power. Finally, the absence of longitudinal data precludes the evaluation of disease progression and the temporal evolution of asymmetry patterns.

In conclusion, this study provides novel insights into imaging and motor asymmetry in GBA- and LRRK2-associated PD and addresses the SOC model within the context of genetic PD. Further studies, ideally incorporating longitudinal data, are warranted to validate and extend these findings. It should be stressed that the group differences observed, albeit highly significant in most cases, are of modest magnitude and cannot be used for categorization of individual patients. Differences in symmetry are relative and not absolute.

## Data Availability

All data produced in the present study are available upon reasonable request to the authors

## Acknowledgements

Statistical analysis codes used to perform the analyses in this article are shared on Zenodo (doi: 10.5281/zenodo.24302063)

PPMI, a public-private partnership, is funded by The Michael J. Fox Foundation for Parkinson’s Research and funding partners, including 4D Pharma, AbbVie, AcureX, Allergan, Amathus Therapeutics, Aligning Science Across Parkinson’s, AskBio, Avid Radio-pharmaceuticals, BIAL, BioArctic, Biogen, Biohaven, BioLegend, BlueRock Therapeutics, Bristol-Myers Squibb, Calico Labs, Capsida Biotherapeutics, Celgene, Cerevel Therapeutics, Coave Therapeutics, DaCapo Brainscience, Denali, Edmond J. Safra Foundation, Eli Lilly, Gain Therapeutics, GE HealthCare, Genentech, GSK, Golub Capital, Handl Therapeutics, Insitro, Jazz Pharmaceuticals, Johnson & Johnson Innovative Medicine, Lundbeck, Merck, Meso Scale Discovery, Mission Therapeutics, Neurocrine Biosciences, Neuron23, Neuropore, Pfizer, Piramal, Prevail Therapeutics, Roche, Sanofi, Servier, Sun Pharma Advanced Research Company, Takeda, Teva, UCB, Vanqua Bio, Verily, Voyager Therapeutics, the Weston Family Foundation, and Yumanity Therapeutics.

